# Monoclonal antibodies against SARS-CoV-2: potential game-changer still underused

**DOI:** 10.1101/2021.09.27.21264191

**Authors:** Ivan Gentile, Alberto Enrico Maraolo, Antonio Riccardo Buonomo, Mariano Nobile, Prisco Piscitelli, Alessandro Miani, Nicola Schiano Moriello

## Abstract

Even several months after the start of a massive vaccination campaign against COVID-19, mortality and hospital admission are still in considerable numbers in many nations. Monoclonal antibodies are the ideal complement to vaccination in high-risk subjects who have been infected by SARS-CoV-2 and are at high risk of developing severe disease. Combining data provided by clinal trials and demographics of SARS-CoV-2 infections, this analysis tries to predict the benefits of an extensive use of monoclonal antibodies to reduce hospital admissions, deaths, and costs.

## Introduction

Vaccination is the most effective way to protect from coronavirus disease 2019 (COVID-19), although it cannot represent the only way to cope with the ongoing pandemic (1–4). Italy is among the Western European countries most hit by the pandemic (5). Notwithstanding campaigns to provide extensive vaccine coverage and information about immunization, part of the population remains sceptical about the vaccination. Several factors, namely safety concerns, doubts about effectiveness, and the speed with which vaccines were developed contribute to vaccine hesitancy and it is unlikely that everyone will eventually agree to vaccination. Moreover, vaccination is less effective in immunosuppressed subjects such as transplanted patients and those with haematological malignancies. A significant proportion of these subjects will not develop an effective response and will be at risk of a severe disease.

Currently, the only effective therapy in patients with mild COVID-19 and who are risk of progression to severe disease is the early (ideally within 4-5 days from symptoms onset) administration of monoclonal antibodies against the spike protein. This therapy reduces both hospitalisations and deaths (6). In the attempt to provide real-life data on the current use of monoclonal antibodies against SARS-CoV-2 in western countries, we investigated the use of monoclonal antibodies in Italy, and estimated the impact of a more extensive use of this therapy on hospital admissions, deaths and costs.

## Methods

We determined the number of new SARS-CoV-2 infections, hospitalisations, ICU admissions and death due to coronavirus disease in Italy between 2 April 2021 (the day Italy approved this therapy) and 5 August 2021. Data of new cases of SARS-CoV-2 infection are publicly available on the website of the Italian Ministry of Health (7). Using WebPlotDigitizer software, we extracted all other data from the graphs on the web page of the Italian National Institute of Nuclear Physics that were provided by Italian National Institute of Health (8). We assumed three scenarios of monoclonal antibody use in people over 70 years of age (rates of use of 50%, 70% or 90%), and estimated their respective consequences on hospital admissions, deaths and costs based on data reported in the REGEN-COV trial (NCT04425629) (6) because complete data were not available for the other approved monoclonal antibodies (i.e. Bamlanivimab plus Etesevimab and Sotrovimab) (9,10). We also estimated the potential economic benefit related to the use of monoclonal antibodies. The price of a monoclonal antibody was estimated at $1,250 per treatment (11). The cost per hospitalisation was estimated as follows: without ICU $16,924, with ICU $57,934 (12). We assumed a ratio between hospitalisation without ICU and with ICU of 9:1, as seen in the target population in the study period, therefore the mean hospitalization cost was set at $ 21,025.

## Results and discussion

As shown in Table 1, 777,502 cases of SARS-CoV-2 infection were recorded during the study period. The incidence fluctuated from a high of 110,594 cases per week to a low of 5,099 cases per week. A total of 70,022 people were over the age of 70 years. There were 57,740 hospital admissions, of which 21,503 in people over 70 years of age. A total of 18,422 deaths were recorded, 9,963 of which among people over the age of 70.

**Table 1.** Data obtained in the period of interest.

|  | Total | Patients over 70 years<br>(rate) |
| --- | --- | --- |
| NEW SARS-COV-2 INFECTIONS | 777,502 | 70,022 (9%) |
| HOSPITAL ADMISSIONS | 57,440 | 21,503 (37.4%) |
| DEATHS | 18,422 | 9,963 (54%) |
| MONOCLONAL ANTIBODIES<br>ADMINISTRED | 6,322 | n.a. |

In Italy monoclonal antibodies use is approved for patients with SARS-CoV-2 infection and with at least one risk factor for progression towards a severe disease. Age over 65 years is one of these factors. In the period considered, monoclonal antibodies were prescribed for 6,322 patients. It is not known how many monoclonal antibodies were prescribed in patients over 70. However, even assuming that all 6,322 treatments were administered in patients in this age group, this figure represents 9.02% of all patients over 70.

Projecting a reduction in hospital admissions and deaths based on the data available (6), we estimate that, during the study period, hospital admissions and deaths would have declined by 13,798 and 6,313 respectively considering a rate of utilization of 90% as shown in Table 2.

**Table 2.** Projected hospital admissions and deaths in subjects above 70 years of age who received monoclonal antibodies.

|  | 50% M.A. USE | 70% M.A. USE | 90% M.A. USE |
| --- | --- | --- | --- |
| **HOSPITAL ADMISSIONS** | 13,837 (15,944 – 12,590) | 10,771 (13,721 – 9,025) | 7,705 (11,498 – 5,460) |
| **REDUCTIONS** | 7,666 (5,559 – 8,913) | 10,732 (7,782 – 12,478) | 13,798 (10,005 – 16,043) |
| **REDUCTIONS (%)** | 35.7% (25.9% - 41.5%) | 49.9% (36.2% - 58.0%) | 64.2% (46.5% - 74.6%) |
| **DEATHS** | 6,456 (8,389 – 5,624) | 5,053 (7,759 – 3,889) | 3,650 (7,130 – 2,153) |
| **REDUCTIONS** | 3,507 (1,574 – 4,339) | 4,910 (2,204 – 6,074) | 6,313 (2,883 – 7,810) |
| **REDUCTIONS (%)** | 35.2% (15.8% - 43,6%) | 49.3% (22.1% - 61.0%) | 63.4% (28.4% - 78.4%) |

M.A.; monoclonal antibodies; in parenthesis data with 95% C.I.

Complete projections are reported in Table 2.

Regarding costs, the expenditure for the acquisition of monoclonal antibodies is counterbalanced by the saving of hospital admissions as a consequence we estimate a total savings of $211,338 million dollars in case of 90% use (see Table 3).

**Table 3:** Economic implications of extensive monoclonal prescription in patients over the age of 70.

| **PERCENTAGE OF M.A. USE** | **50%** | **70%** | **90%** |
| --- | --- | --- | --- |
| **NUMBER OF A.M. ADMINISTERED** | 35,011 | 49,015 | 63,020 |
| **COST OF M.A. (\$)** | 43,763,750 | 61,269,250 | 78,774,750 |
| **SAVING FOR H.A. REDUCTION (\$)** | 161,173,855 | 225,643,397 | 290,112,939 |
| **NET SAVING (\$)** | 117,410,105 | 164,374,147 | 211,338,189 |

H.A.: Hospital Admissions; M.A. monoclonal antibodies

We acknowledge that this study presents several limitations; first of all, we projected the efficacy of monoclonal antibodies estimating a similar effectiveness in real-life compared to that from clinical trials. For example, the population considered in the present study was significantly different from that one enrolled in the trials at least in terms of median age; moreover, the estimate of the use of monoclonal antibodies is rough because it does not consider other categories in which monoclonal antibodies are indicated (e.g., subjects with immunodeficiency, diabetes, obese, etc.)

## Conclusion

Use of monoclonal antibodies for SARS-CoV-2 infection between April and August 2021 in Italy has been very low. Even looking at the most prudential estimates, a more extensive use of these therapies could have prevented a high number of hospitalisations and deaths among patients over 70, together with a positive impact on COVID-19 related costs. These data should prompt authorities to create a higher interaction between general practitioners and facilities for monoclonal antibodies infusion, in order to refer patients at high-risk of deterioration as soon as possible.

## Data Availability

All data utilized in this article are publically available.

